# A translational multimodal machine-learning prototype predicting valproate response in epilepsy treatment

**DOI:** 10.1101/2025.08.21.25332294

**Authors:** Simeon Platte, Afsheen Kumar, Giorgia Guerini, Massimo Pandolfo, Denise Haslinger, Colin B. Josephson, Guillermo Delgado-García, Navprabhjot Kaur, Michaela-Pauline Lux, Heiko Stempfle, Chantal Depondt, Reetta Kälviäinen, Felix Rosenow, Sophie von Brauchitsch, Karl Martin Klein, Andreas G. Chiocchetti

**Author notes:** **Funding:** RAISE-GENIC; EU-AIMS2-Trials, R2D2-MH.

## Abstract

Epilepsy affects around 1% of the global population and often requires long-term treatment with antiseizure medications (ASMs). However, the current treatment strategy is based on clinical acumen and trial and error, resulting in only about 50% of patients remaining seizure-free for at least 12 months with first-line ASMs. Valproic acid (VPA) is a commonly prescribed first-line ASM, yet <50% of patients experience inadequate seizure control (ISC) or unacceptable adverse reactions (UARs), necessitating discontinuation. We developed a predictive algorithm to support VPA treatment decisions in eligible epilepsy patients integrating in-vitro data, genetic data and previous knowledge. Our approach is based on genetic variations in genes associated with VPA pharmacodynamics and -kinetics, as well as the response of human neurons to VPA (*in-vitro*). The multimodal pipeline integrates patients’ common and rare genetic variants of genes related to these pathways. The feature engineering pipeline was trained and tested on a multi-ethnic external dataset and the final classifier trained and tested on the Epi25 cohort of patients. The proof-of-concept validation was performed in an independently collected cohort and confirmed the potential to predict VPA treatment response. Overall performance was modest. However, prediction accuracy and the high negative predictive value highlight the potential for clinical values. We estimated a significant reduction in the time to successful treatment, decreasing both patient burden and overall healthcare costs. While our prototype is not yet at a clinically-ready stage and the need for SNP and WES data is limiting feasibility, the results suggest that a translational biomarker-based algorithm is promising in personalizing epilepsy treatment with VPA, shifting away from the one-size-fits-all approach. This could enhance treatment efficacy, reduce ISC and UARs, and improve patients’ quality of life by shortening the time to achieve seizure freedom.

## Introduction

Epilepsy is a chronic neurological disorder affecting approximately 1% of the global population^1^. It is characterized by recurrent seizures and in most cases requires long-term treatment with antiseizure medications (ASMs). However, the current strategy for identifying the best treatment is based on solely on clinically guided trial-and-error methods.. Only around 50% of patients remain seizure-free for at least 12 months with first-line drug treatment, 12% with the 2^nd^ and 4% with the 3^rd^ treatment regim^2–4^; 33% of patients exhibit incomplete response to ASMs ^3,5^. The median time to failure ^6^ is 90 days due to unacceptable adverse reactions (UARs) and 234 days due to inadequate seizure control (ISC).. A treatment decision informed by multimodal data could substantially reduce the time required to achieve seizure freedom. Based on the current accuracy and median time to failure, a treatment decision support tool performing with only 60% accuracy would be able to reduce the average time to successful treatment by 19.41 days per patient and any increase by one percent reduces the delay by an additional 2.04 days. A reduction in time to failure reduces patient burden, improves quality of life (QoL), medication adherence^7^ reduces the staffing requirements and thus overall costs ^3^. Although many new ASMs have been introduced in the last years, there has been little improvement in overall treatment success rates ^4,6^. This underpins the need for personalized or subgroup-specific treatments, rather than a one-size-fits-all approach ^3^. Thus, a personalized approach to epilepsy treatment is key ^7^.

One of the most prescribed first-line ASMs, consistently since its approval in 1967, is Valproic acid (VPA) in children and adolescents (29.6%) as well as adults (26.5%) ^8^, though its use in females has declined likely due to wider recognition of its teratogenic effects ^9^.

However, 40-50% of patients experience ISC or UARs and must discontinue the medication (37% in SANAD I & II generalized or unclassifiable epilepsy trials ^4,6^). The main VPA UARs include hepatotoxicity, pancreatitis, weight gain, irregular periods, cognitive difficulties in particular with spatial memory, alopecia, tremor, osteoporosis, and teratogenicity. Several mechanisms have been hypothesized to be associated specifically with UAR of VPA^10^ (39871487) such as mitochondrial dysfunction, carnitine shortage, immune-mediated reactions, glutathione depletion or leptin and insuline resistance^11^. However the pathophysiologically relevant genes and variations are not as well defined as VPAs pharmacokinetic and pharmacodynamic pathways.

We here developed a predictive, biomarker-based algorithm to support the treatment decision for VPA in epilepsy.

Our approach is based on two main assumptions: i) genetic variation in genes associated with VPA pharmacodynamics and pharmacokinetics, as well as ii) cellular response to VPA, are potentially stable predictors of treatment response.

VPA is metabolized via glucuronidation, β-oxidation in the mitochondria, and to a lesser extent via cytochrome P450 (CYP)-mediated oxidation ^12^. At the functional level, VPA has been identified to act on γ amino butyric acid (GABA) levels in the brain by blocking the major anabolic enzymes 4-Aminobutyrate Aminotransferase (ABAT), Aldehyde Dehydrogenase 5 family member A1 (ADH5A1) as well as the oxoglutarate dehydrogenase (OGDH) (reviewed in ^13^). VPA further blocks voltage-gated ion channels CACNA1C, CACNA1D, CACNA1N, CACNA1F and the SCN family. In addition, it has been reported to inhibit the Histone Acetylase HDAC1 and activates HDAC9 and HDAC1 ^14,15^, increasing the expression of antiapoptotic and antitumor genes.

We further here extended this list of genes including those regulated upon response to VPA in human GABAergic and glutamatergic neurons, as assessed in an *in-vitro* model.

In this study, which is part of the EU-funded RAISE-GENIC consortium, we developed a multimodal machine-learning pipeline on features derived from an individual’s genetic variants of genes associated with VPA metabolism, function and gene-network regulation and tested if we can predict the response to treatment. The chosen model architecture has the potential to scale to other ASMs or phenotypes where physiological mechanisms are known.

This prototype was developed together with patient representatives and was trained and tested in a multi-ethnic cohort and validated in an independently collected sample.

## Methods

We built a multimodal machine learning pipeline (Figure 1) to predict valproic acid (VPA) response using genetic and clinical data. The model was trained and tested on a Discovery cohort (N=329, Epi25) and finally validated on an independent cohort (N=202, newly recruited). Key parameters are summarized below; full methods are in the Supplementary Material.

**Figure 1:**
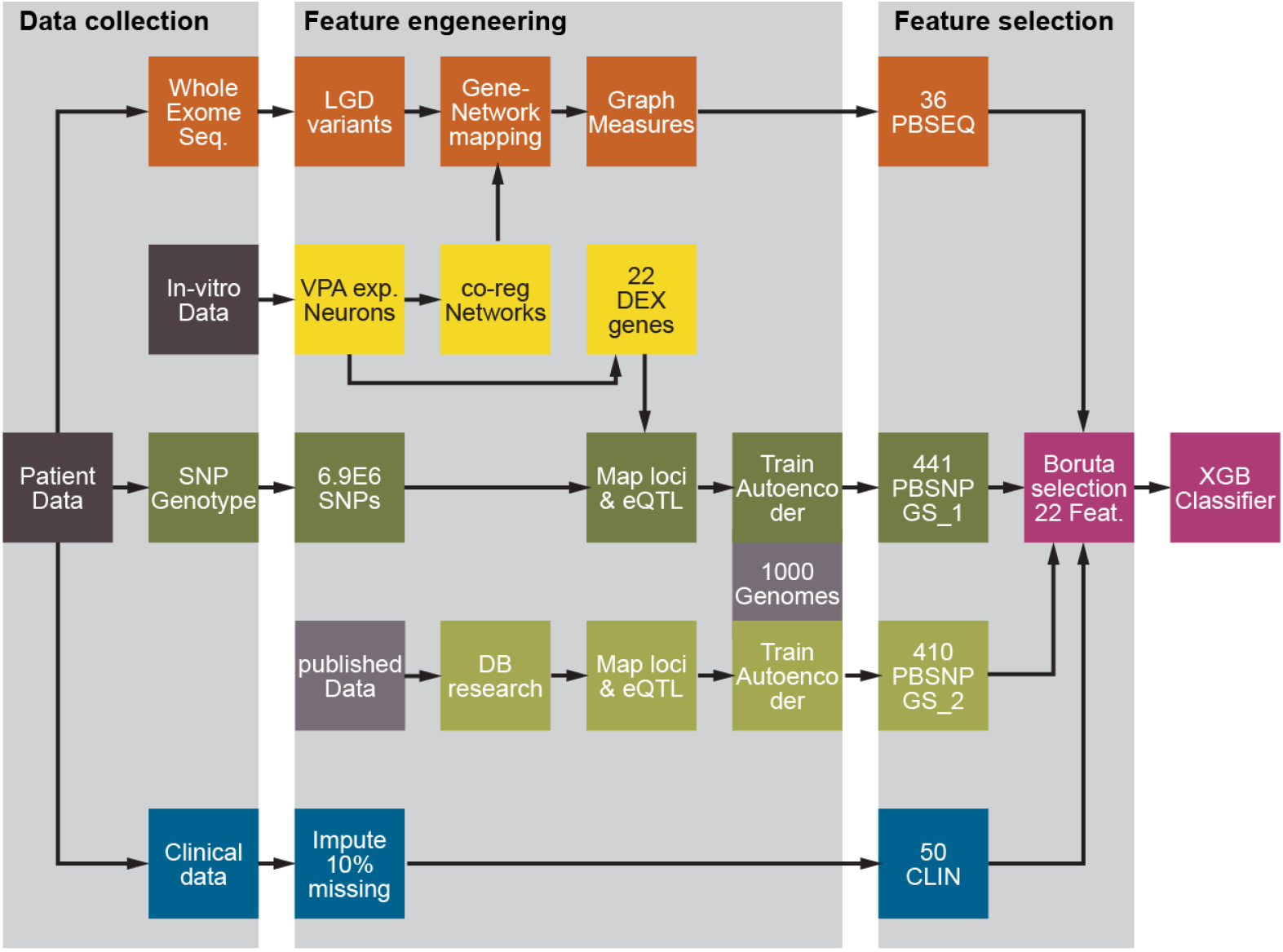
Schematic overview of the implemented multimodal pipeline. The final Classifier (purple) integrated three different feature sets: Network-based features derived from Whole Exome Sequencing (PBSEQ), Common Variation derived features of Valproic Acid (VPA) associated pathways (PBSNP) from two Gene Sets (GS_1, GS_2). Newly generated data (dark gray) and published data (light gray) were included to define these Gene Sets. The pipeline was implemented and trained on a Train-Discovery cohort. Parameters and Models were then tested in the Test-Discovery cohort and finally validated in an independent Validation cohort. CLIN: Clinical Features; DB: Database; DEX genes: Differentially Expressed Genes; eQTL: Expression Quantitative Trait Loci; GS: Gene Set; LGD: Likely Gene-Disrupting (variants); PBSEQ: Pathway-Based Sequencing Features; PBSN: Pathway-Based SNPs; VPA exp. Neurons: Valproic Acid Exposed Neurons; XGB Classifier: eXtreme Gradient Boosting Classifier (XGBoost)

### Definition of gene sets

#### Gene Set 1: Valproate associated genes and gene-networks

Human induced pluripotent stem cells (iPSCs) derived from two healthy donors were differentiated to cortical progenitors and neurons for 40 days following established protocols.^16^. On day 35, cells were exposed to 0.0, 0.33 and 1.0 mM VPA respectively, based on previous experiments^17^ (Supplementary Figure S 1). On day 40 RNA was extracted and 3′ RNA sequencing (single end, two replicates per condition; N=12 total) was performed. The RNA analysis yielded 15,629 genes after quality control; no sample was excluded due to technical outliers (Supplementary Figure S 2). Differential gene expression analysis in response to VPA was performed using linear mixed models and the DESeq2 pipeline ^18^, following established methods as previously published ^19^. Differentially expressed genes (DEGs) replicated in both donor cell lines (FDR < 0.05, |log_2_FC| > 1) formed **Gene Set 1**. To identify VPA-responsive gene modules, we applied weighted gene co-expression network analysis (WGCNA ^20^). Differentially expressed Modules replicated with FDR < 0.05 and |log_2_FC| > 1 were included.

#### Gene Set 2 VPA Pharmacogenomics pathways

To account for pharmakokinetic, pharmacodymic effects as well as related adverse responses (UARs), Gene Set 2 comprised VPA metabolism-associated genes (cytochrome P450 and UDP-glucuronosyltransferases) from DrugBank (ID: DB00313, accessed 2023-09-14).

### Data description

#### PBSNP embedding cohort

SNP-based autoencoders were trained on 3,157 samples from the 1000 Genomes Phase 3 dataset ^21^, filtered to 4.1Mio SNPs shared with the Discovery cohort.

#### Treatment Response Labels

Among the 1,334 individuals included, 417 received VPA as a monotherapy, either as a first-line, second-line, or subsequent treatment at any given timepoint. Treatment outcomes were manually classified as Responder, Non-responder, Unclassified, or Unknown per site. Responders achieved ≥12 months seizure freedom attributed to VPA. Non-responders had >50% seizure recurrence or discontinued VPA (due to inefficacy, adverse effects, or unknown reasons). Unclassified had partial response or unclear data. Unknown cases had no information on treatment response documented and were excluded. Following clinical and patient advisory input, non-responders and unclassified were grouped, yielding 156 Responders and 173 non-Responders.

#### Discovery cohort

Data from 1,334 epilepsy patients across three Epi25 ^22,23^ sites (Brussels N= 387, Frankfurt N= 259 and Kuopio N= 688) were included. After QC and imputation as previously published ^24^, 1,327 individuals with 4.1 M high-quality SNPs remained. Whole exome sequencing (WES) was available for 1,319 individuals; likely gene-disrupting (LGD) variants were identified using ANNOVAR ^25^. Clinical data was available for 1,244 patients, including seizure/epilepsy type (for a full list of features, see Supplementary Table S 2). After excluding individuals with >10% missing clinical data, remaining gaps were imputed (MICE ^26^ or zero-imputation of sparse binary features).

#### Independent Validation Cohort

The independently recruited cohort of 202 epilepsy patients from Calgary, included SNP genotyping for 168 individuals. Preprocessing was performed as above. Coverage of Gene Set 1 and 2 SNPs was >94%. Missing SNPs were imputed using major alleles. WES data was available for 167 individuals and processed as described above.

Clinical variables matched the Discovery cohort and were available for 148 patients. Sparse binary features were zero-imputed; age of onset was MICE-imputed (3.4% missing). After excluding 19 patients with unknown VPA outcomes, 49 responders, 134 non-responders remained.

### Feature Engineering

#### PBSNP feature engineering

To capture diverse genetic backgrounds, one undercomplete autoencoders (AE) per Gene Set was trained on the 1000 Genomes dataset (80/20 train-test split). SNPs within ±2kb of genes in Sets 1 and 2, plus GTEx v8 brain eQTLs ^27^, were included. Variants in linkage disequilibrium (R^2^ > 0.8) were pruned, yielding 4,093 SNPs (Gene Set 1) and 27,870 SNPs (Gene Set 2). Minor allele dosages were scaled to 0, 0.5, and 1. AEs used the Adam optimizer, LeakyReLU activation, and MSE loss. Bottleneck size was set to 10% of input size; batch size was 32. A grid search (36 models; Supplementary Table S **3**) varied dropout rates, activation functions, and network depth. Models were trained for up to 50 epochs with early stopping and evaluated via 3-fold cross-validation. Final models were retrained for up to 500 epochs and validated on the 20% hold-out set. Bottleneck layer outputs from the best models were used as compressed PBSNP features for each patient.

#### PBSEQ: Network-Based Features from Exome Data

PBSEQ features captured graph-based properties of the VPA-responsive gene modules. For each module, 12 graph metrics were computed from the gene-gene correlation matrix, including weighted (e.g., mean/sum connectivity, path length, skewness) and unweighted (e.g., density, betweenness, transitivity) parameters. For unweighted measures, edge correlations were thresholded preserving 95% of node connectivity. To calculate patient-specific features, genes with likely gene-disrupting (LGD) variants were removed from the graph, and network metrics were recalculated.

### Multimodal Data Integration

#### Discovery cohort

Features from three datasets i.e. PBSNP (Gene Sets 1 & 2), PBSEQ, and CLIN (Clinical data) were combined for 329 patients with VPA outcome data. Complete multimodal data is available for 308 patients.

### Validation cohort

Integration followed the same approach, yielding 156 individuals with complete multimodal data.

### Classifier / Recommender Model

The Discovery cohort was split into a stratified training set (60%) and hold-out test set (40%), preserving responder ratios (47%). Synthetic Minority Oversampling Technique (SMOTE^28^) was applied independently to balance classes, generating 17 synthetic responders per set. Features were min-max scaled on the training set and applied to test and validation sets.

#### Feature selection

Features were selected using the Boruta ^29^ algorithm (p > 0.01, max 200 runs). The union of features from 10 runs was used to maximize relevant and tentative predictors.

#### Model training

A gradient-boosted decision tree classifier (XGBoost ^30^) was trained on selected features to predict VPA response. Hyperparameters were tuned via cross-validated grid search (early stopping = 20, max 1,000 rounds; see Supplementary Table S 4). The best model was retrained with optimized parameters (early stopping = 30, max 5,000 rounds).

#### Validation

Model performance was assessed on the hold-out test set and validated on the independent validation cohort using the full pipeline.

#### Model comparison

To assess modality contribution, six additional models using individual or paired feature types (PBSNP, PBSEQ, CLIN) were trained and validated using the same full pipeline.

## Data availability

All software used is open source (Python 3.10 and in R 4.3.3). The full code is publicly available under https://gitlab.rz.uni-frankfurt.de/cap_molgenlab/epi-vpa-ml-pipeline. Pre-trained models and weights as well as patient data are available upon request.

## Ethics statement

Informed consent for the EPi25 Collaborative is documented here^22^. The recruitment of the Validation Cohort in Canada was approved by the Conjoint Health Research Ethics Board at the University of Calgary (REB18-2099). Collection and generation of iPSCs was done according to a protocol approved by the institutional Ethics Committee of Erasmus Hospital, Brussels (P2008.313). All participating individuals provided written informed consent.

## Results

### Sample characteristics: Responders vs non-responders

The overall Discovery cohort included 329 individuals with genetic and clinical data available. Significant differences (uncor. p <0.05) between VPA responders and non-responders were observed (Table 1). As expected, non-responders showed higher rates of drug resistance (variable not included in final classifier), regional epileptiform discharges (focal) discharges, focal seizures, and bilateral seizures (OR > 1), and lower rates of photoparoxysmal reactions, generalized seizures, and myoclonic seizures. No significant differences were found for age at onset, age at study, or sex. After stratified randomization, the cohort was split into training (60%) and test (40%) sets. The only difference between sets was a higher proportion of females in the test set (p=0.042; Supplementary Table S 5).

**Table 1:** Descriptive Statistics Discovery cohort.

|  | Responders<br>N=156 | Non-Responders<br>N=173 | OR [95CI] | p<sup>a</sup> |
| --- | --- | --- | --- | --- |
| Cohort: |  |  |  |  |
| Test-disc | 63 (40.385%) | 70 (40.462%) |  |  |
| Train-disc | 93 (59.615%) | 103 (59.538%) | 1.00 [0.64;1.55] | 1.000 |
| Sex: |  |  |  |  |
| Female | 91 (58.333%) | 100 (57.803%) |  |  |
| Male | 65 (41.667%) | 73 (42.197%) | 1.02 [0.66;1.59] | 1.000 |
| Enrolment age | 44.052 (17.878) | 47.846 (17.712) |  | 0.062 |
| Age onset | 21.647 (17.465) | 20.580 (16.052) |  | 0.583 |
| Ethnicity PC1 | 0.020 (0.018) | 0.016 (0.023) |  | 0.053 |
| Ethnicity PC2 | -0.004 (0.011) | -0.005 (0.033) |  | 0.780 |
| Ethnicity PC3 | 0.001 (0.011) | -0.003 (0.046) |  | 0.212 |
| Ethnicity PC4 | -0.001 (0.011) | 0.001 (0.033) |  | 0.440 |
| Autism: | 0 (0%) | 0 (0%) |  |  |
| Intellectual disability: | 10 (6.410%) | 12 (6.936%) | 1.09 [0.45;2.67] | 1.000 |
| Psychosis: | 6 (3.846%) | 10 (5.780%) | 1.52 [0.54;4.64] | 0.577 |
| Spasms seizures: | 0 (0%) | 0 (0%) |  |  |
| Focal epileptiform discharges: | 49 (31.410%) | 88 (50.867%) | 2.25 [1.44;3.56] | 0.001 |
| Photoparox. react: | 5 (3.205%) | 0 (0.000%) | Inf | 0.023 |
| Epileptiform unspec: | 8 (5.128%) | 12 (6.936%) | 1.37 [0.54;3.63] | 0.650 |
| Focal seizures: | 90 (57.692%) | 149 (87.135%) | 4.92 [2.88;8.69] | <0.001 |
| Fever related seizures: | 15 (9.615%) | 24 (13.873%) | 1.51 [0.76;3.06] | 0.307 |
| Bilateral seizures: | 70 (45.161%) | 106 (61.272%) | 1.92 [1.24;2.99] | 0.005 |
| Generalized seizures: | 48 (30.968%) | 11 (6.395%) | 0.15 [0.07;0.30] | <0.001 |
| Tonic seizures: | 0 (0.000%) | 1 (0.578%) | . [.;.] | 1.000 |
| Atonic seizures: | 3 (1.935%) | 2 (1.156%) | 0.61 [0.07;4.04] | 0.670 |
| Myoclonic seizures: | 24 (15.385%) | 8 (4.651%) | 0.27 [0.11;0.61] | 0.002 |
| Absence seizures: | 24 (15.385%) | 5 (2.890%) | 0.17 [0.05;0.42] | <0.001 |
| Atypical absence sz.: | 2 (1.333%) | 2 (1.176%) | 0.88 [0.09;8.55] | 1.000 |
| Other seizures: | 1 (0.641%) | 2 (1.176%) | 1.73 [0.14;54.8] | 1.000 |

a) Uncorrected test of significance: Chi<sup>2</sup> for dichotomous and t-test for continuous variables. Inf:infinity, sz: seizures, unspec: unspecified

The Validation cohort showed a similar clinical profile to the Discovery cohort (Table 2). Non-responders had higher rates of focal discharges, focal and bilateral seizures, and lower rates of photoparoxysmal reactions, generalized, and myoclonic seizures. No differences were observed in sex, age at onset, or age at study. Drug resistance data was unavailable, and non-responders showed a higher prevalence of intellectual disability.

**Table 2:**
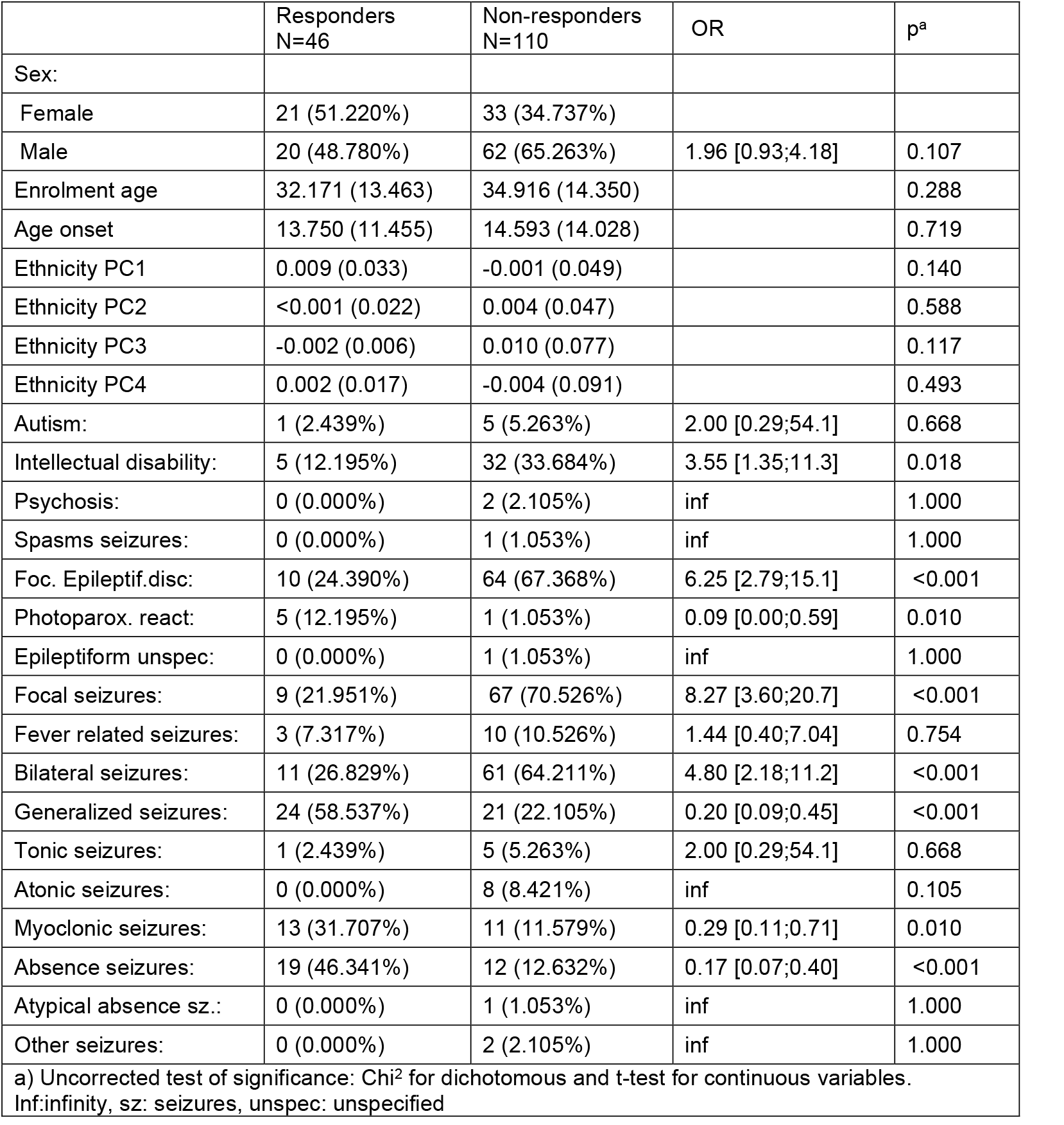
Descriptive Statistics Validation cohort.

|  | Responders<br>N=46 | Non-responders<br>N=110 | OR | p<sup>a</sup> |
| --- | --- | --- | --- | --- |
| Sex: |  |  |  |  |
| Female | 21 (51.220%) | 33 (34.737%) |  |  |
| Male | 20 (48.780%) | 62 (65.263%) | 1.96 [0.93;4.18] | 0.107 |
| Enrolment age | 32.171 (13.463) | 34.916 (14.350) |  | 0.288 |
| Age onset | 13.750 (11.455) | 14.593 (14.028) |  | 0.719 |
| Ethnicity PC1 | 0.009 (0.033) | -0.001 (0.049) |  | 0.140 |
| Ethnicity PC2 | <0.001 (0.022) | 0.004 (0.047) |  | 0.588 |
| Ethnicity PC3 | -0.002 (0.006) | 0.010 (0.077) |  | 0.117 |
| Ethnicity PC4 | 0.002 (0.017) | -0.004 (0.091) |  | 0.493 |
| Autism: | 1 (2.439%) | 5 (5.263%) | 2.00 [0.29;54.1] | 0.668 |
| Intellectual disability: | 5 (12.195%) | 32 (33.684%) | 3.55 [1.35;11.3] | 0.018 |
| Psychosis: | 0 (0.000%) | 2 (2.105%) | inf | 1.000 |
| Spasms seizures: | 0 (0.000%) | 1 (1.053%) | inf | 1.000 |
| Foc. Epileptif.disc: | 10 (24.390%) | 64 (67.368%) | 6.25 [2.79;15.1] | <0.001 |
| Photoparox. react: | 5 (12.195%) | 1 (1.053%) | 0.09 [0.00;0.59] | 0.010 |
| Epileptiform unspec: | 0 (0.000%) | 1 (1.053%) | inf | 1.000 |
| Focal seizures: | 9 (21.951%) | 67 (70.526%) | 8.27 [3.60;20.7] | <0.001 |
| Fever related seizures: | 3 (7.317%) | 10 (10.526%) | 1.44 [0.40;7.04] | 0.754 |
| Bilateral seizures: | 11 (26.829%) | 61 (64.211%) | 4.80 [2.18;11.2] | <0.001 |
| Generalized seizures: | 24 (58.537%) | 21 (22.105%) | 0.20 [0.09;0.45] | <0.001 |
| Tonic seizures: | 1 (2.439%) | 5 (5.263%) | 2.00 [0.29;54.1] | 0.668 |
| Atonic seizures: | 0 (0.000%) | 8 (8.421%) | inf | 0.105 |
| Myoclonic seizures: | 13 (31.707%) | 11 (11.579%) | 0.29 [0.11;0.71] | 0.010 |
| Absence seizures: | 19 (46.341%) | 12 (12.632%) | 0.17 [0.07;0.40] | <0.001 |
| Atypical absence sz.: | 0 (0.000%) | 1 (1.053%) | inf | 1.000 |
| Other seizures: | 0 (0.000%) | 2 (2.105%) | inf | 1.000 |
| a) Uncorrected test of significance: Chi<sup>2</sup> for dichotomous and t-test for continuous variables.<br>Inf:infinity, sz: seizures, unspec: unspecified |  |  |  |  |

### iPSC results and definition of Gene Set 1 and Gene networks

After 40 days of differentiation, iPSC-derived neurons expressed β3-tubulin, VGLUT-1, GAD65, and S100β, confirming neuronal identity (Figure 2 A-B). Cell viability was assessed after VPA exposure from 0.0625-1.0 mM (Figure 2 C) and thus the clinical concentration of 0.33 and 1mM were used. Differential expression analysis identified 22 genes significantly associated with VPA concentration (0, 0.33 and 1.0 mM) in both biological replicates (Figure 2 D), including *ATP1A2* (*log_2_FC* = –1.836, *FDR* = 0.014) and *KCNA1* (*log_2_FC* = 2.620, *FDR* = 0.043), both linked to epilepsy ^31,32^. The coding regions of these genes (±2kb) and their eQTLs yielded 4,093 SNPs used for PBSNP feature engineering (see below).

**Figure 2:**
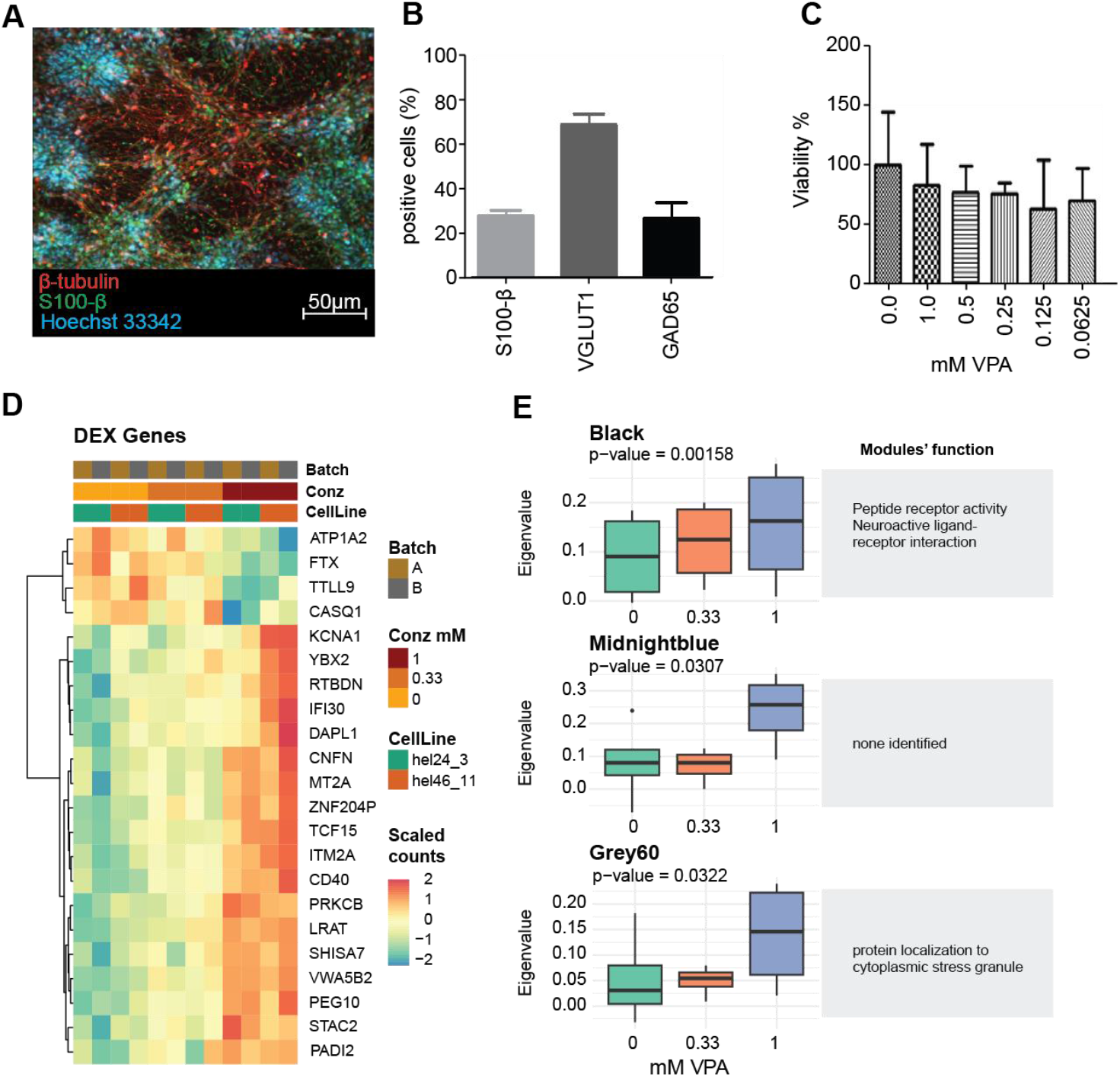
Identification of Genes and gene network differentially expressed in iPSC derived Neurons. A) Representative IHC image of differentiated neurons. Cortical neurons were stained with the neuronal marker β-tubulin (red) and glia cells with the S100-b marker. Nuclei are Hoechst 33342 positive (blue) B) Quantification of glia cells (Sβ100) as well as glutamatergic (VGLUT1) and GABAergic (GAD65) neurons based on IHC-staining. Proportions refer to Hoechst-stained nuclei. C) Viability assay determining the optimal concentration of VPA. Since the physiologically reported concentration of 1mM and 0.33 mM showed acceptable viability, these were chosen for treatment. Percentages refer to the mean Viability at 0mM. D) Heatmap of DEX genes upon valproate treatment. Only genes with a |log2FC|>1 and an FDR<0.05 were considered. E) Modules identified in the WGCNA with a differential Eigengene Value expression upon VPA treatment (|log2FC|>1 and an FDR<0.05). All three modules were up-regulated. Results of Gene Ontology (GO)-term enrichment analysis are displayed below the modules.

WGCNA identified three VPA-upregulated co-expression networks (Figure 2 E). i) Black module (595 genes), associated with Neuroactive ligand-receptor interaction (p=0.026), ii) Midnight module (269 genes; function unknown), iii) Gray60 module (243 genes) associated with localization to cytoplasmic stress granule (p=9E-4). These networks formed the basis for PBSEQ graph feature generation.

### Feature engineering & generation

#### PBSNP autoencoder training results and feature extraction

Autoencoders trained on the 1000 Genomes dataset yielded 410 features (Gene Set 1) and 441 features (Gene Set 2), with low reconstruction error (RMSE: 0.217 and 0.267, respectively). Key architectural differences included bottleneck activation and input dropout (Supplementary Table S 3). PBSNP features showed minimal correlation with population structure (SNP based PC1-PC5; mean ***r*** = −0.010, SD = 0.094).

#### PBSEQ Graph Network feature computation

Exome sequencing identified on average 1,120 functional variants (including rare and common SNVs) per individual. Network analysis produced 36 graph-based PBSEQ features with inter-individual variance above zero.

### Feature selection results

Boruta feature selection identified 22 relevant features: 16 PBSNPs (14 from Gene Set 1, 2 from Gene Set 2), 2 PBSEQ metrics (sum connectivity, mean betweenness in gray60 module), and 3 clinical features (focal seizures, generalized seizures, structural MRI findings).

### Classifier training and testing

A gradient-boosted tree model (XGBoost) was trained on the 22 selected features. The best model (subSample = 1, colSample = 0.426, maxDepth = 3, minChildweight: 1, eta = 0.08, trained on the rmse metric and a binary:logistic objective) reached a train-RMSLE of 0.105 (158 iterations) and test-RMSLE of 0.417. In the test set, performance (AUC, F1, Sensitivity and Specificity) exceeded chance with an AUC = 0.66 (Figure 3 A, Supplementary Table S 6).

**Figure 3:**
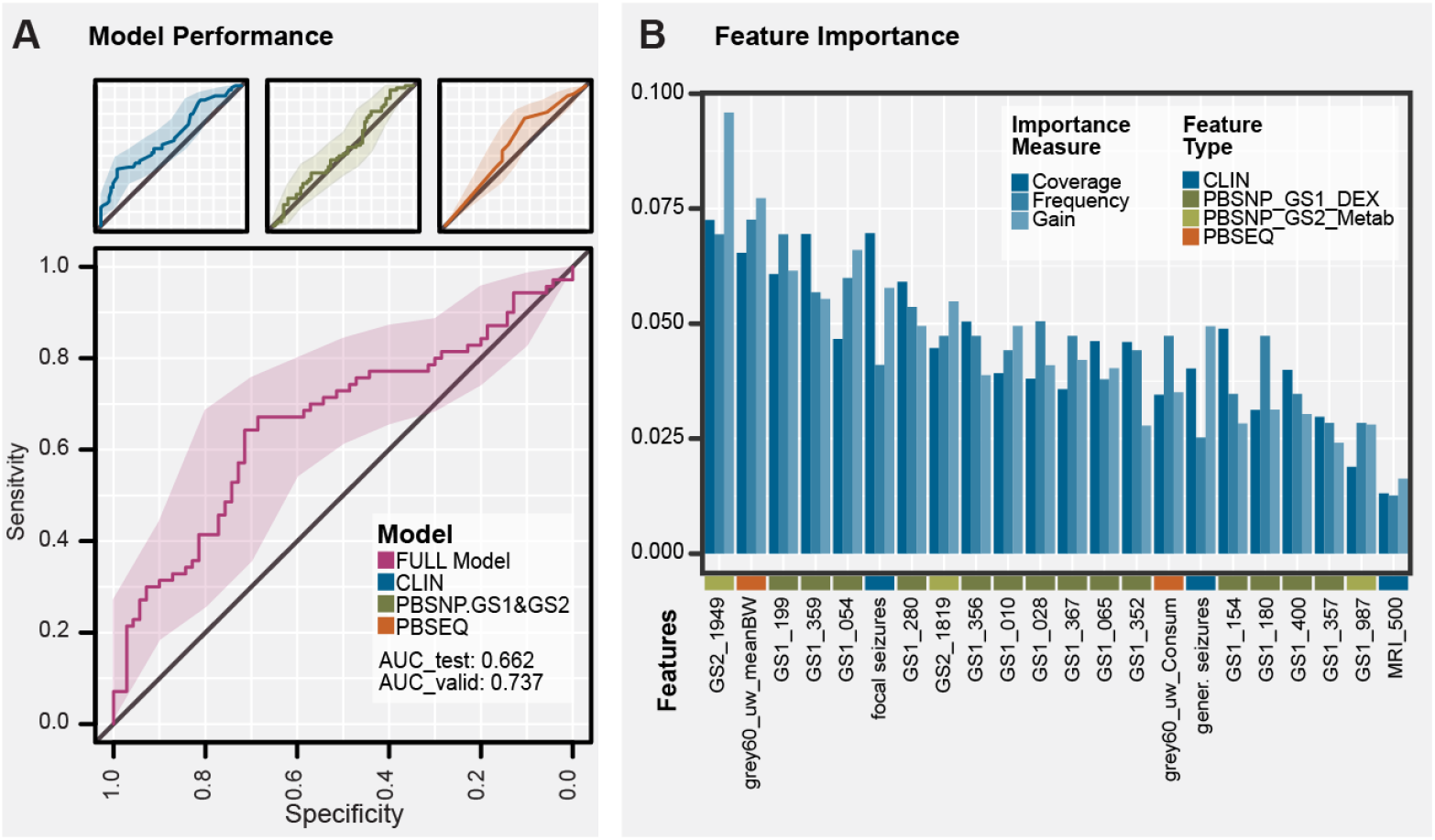
Model Performance and Feature Importance. A) We compared performance of models including different feature sets. The best model included all three types of features (FULL Model). None of the other models achieved a comparable accuray when including only Clinical data (CLIN), PBSNP or PBSEQ features, respectively. For a full list of perfomance metrics see Supplementary Table S 6. The AUC_test refers to the unseen test-Discovery cohort, and the AUC_valid to the newly recruited Validation cohort. B) The full model included a total of 22 features, i.e., 16 PBSNP features from Gene Set 1 (GS1, Differentially Expressed DEX Genes, olive green), 3 from Gene Set 2 (GS2, Metabolism and pharmacodynamics/-kinetics, light green), 2 PBSEQ Network features (orange), and three clinical phenotypes (blue). Features are sorted based on the average of the three importance measures implemented.

Top predictors (Figure 3 B) were primarily PBSNP features from Gene Set 1. However, the most important individual features included one from Gene Set 2 and a PBSEQ feature (mean betweenness, gray60 module).

To assess potential bias, we compared clinical variables between true positives (TP) and false negatives (FN), and between true negatives (TN) and false positives (FP). No significant differences were found in age, sex, or ethnicity, suggesting consistent model performance across demographic groups.

However, focal seizures were significantly less common in TP (26.3%) vs. FN (92.0%), and drug resistance was lower in TP (26.3%) vs. FN (44.0%) (p < 0.001; Supplementary Table S 7). No significant differences were observed between TN and FP groups.

To assess the contribution of each data modality, six models using individual or paired feature sets (PBSNP, PBSEQ, CLIN) were compared to the full model. None outperformed the full model in AUC, sensitivity, or specificity (Supplementary Table S 6). The CLIN-only model had the second-highest balanced accuracy (0.565) but showed lower F1-score (0.559) and AUC (0.640).

The full model was thus selected for external validation. In the independent Validation cohort, it achieved robust performance with an AUC of 0.74 (0.73 balanced AUC), sensitivity of 0.80 (bal. sensitivity = 0.8), and specificity of 0.50 (balanced specificity: 0.46), indicating performance above chance and adaptability of the threshold to clinical goals.

In conclusion, the model performs consistently across demographic groups, though further evaluation is needed to confirm generalizability across diverse clinical settings.

## Discussion

In this study, we developed and validated a machine learning-based decision-support tool to predict valproic acid (VPA) treatment response across diverse epilepsy types. By integrating genetic, transcriptomic, and clinical data, the model achieved a 59–66% prediction accuracy – outperforming clinically guided trial-and-error methods in representative cohorts ^4,33^. This is notable given that success rates typically decline with each subsequent ASM trial, and only ~66% of patients achieve seizure freedom with appropriate therapy ^5^. Our model offers a one-step prediction strategy comparable to the effectiveness of up to three sequential ASM trials.

### Suitability and Scalability of the Algorithm

Assuming a 66% overall ASM response rate and published VPA success rates across treatment lines (50.5%, 11.6%, 4.1%)^3^ with a median 162-day time-to-failure per trial ^6^, current trial-and-error strategies result in an average delay of 32.1 days before reaching an effective treatment. Using our model with a 59% first-line accuracy reduces this delay to 14.7 days: a 17.4-day reduction (54.1%). These estimates are conservative, excluding costs of false positives; full assumptions are detailed in Supplementary Table S8.

The model’s 0.80 sensitivity in the independent validation cohort highlights its strength in correctly identifying VPA responders, potentially reducing ineffective ASM trials. Although specificity was lower (0.50), the model functions effectively as a screening tool, prioritizing sensitivity to avoid missing potential responders. Despite a modest positive predictive value (0.402), the high negative predictive value (0.859) supports its clinical utility in identifying patients unlikely to benefit from VPA.

The model relies on SNP and WES data, which are not yet standard in clinical practice thus limiting feasibility in standard clinical care. For its use to be justified, the predicted time gain must outweigh delays from genetic assessment. With an average gain of 17.4 days, and substantially more in some cases (e.g., 234 days median delay due to ISC ^4,6^), this appears feasible if genetic data can be provided e.g. within one week, or are already available. While upfront costs exist, faster treatment decisions may offset them, though dedicated health economic evaluations are needed.

Technically, the model uses scalable methods, i.e., autoencoder-based feature reduction and gradient-boosted trees, to manage high-dimensional data efficiently. Its reliance on public resources (e.g., 1000 Genomes, GTEx) and standard tools supports adaptability across clinical settings. PBSNP and PBSEQ features improved prediction beyond clinical data, highlighting the value of gene set–based encodings and network-derived features. These approaches may be broadly applicable to other genetically complex traits as well as to other ASMs. Clinical feasibility, however, will depend on local access to rapid, cost-effective genotyping.

Finally, the training, test and validation cohorts where mainly of Western/European Ancestry and thus generalizability across divers genetic population strata needs to be further evaluated. However, post-hoc analysis of false Negatives and False positives was promising, as we did not identify any association with the genetic population stratification dimensions.

### Comparison to Similar Approaches

Prior studies using clinical or genetic predictors for ASM response often relied on single-variable models or small datasets, limiting generalizability. One multimodal study on brivaracetam ^34^ used SNV counts per gene set but did not capture non-linear allele interactions. A deep learning model using 16 clinical variables ^35^ achieved moderate performance (AUROC 0.52–0.65), comparable to our CLIN-only model, underscoring both the feasibility of clinical prediction and the added value of integrating genetic and transcriptomic data.

### Conclusion and Outlook

In summary, our machine learning model provides a scalable tool for predicting VPA response, matching the effectiveness of two to three clinician-guided ASM trials. Early identification of non-responders may reduce adverse effects, accelerate seizure control, and improve outcomes. Future work will extend the model to other ASMs, incorporate additional omics data, and support clinical implementation via a user-friendly interface.

## Data Availability

All data produced in the present study are available upon reasonable request to the authors.

https://gitlab.rz.uni-frankfurt.de/cap_molgenlab/epi-vpa-ml-pipeline

## Acknowledgements

This work was mainly funded by the ERA PerMed Joint Transnational Call 2018 (JTC 2018), co-funded by the European Commission and national/regional funding partners, Project ID: RAISE-GENIC. Partial funding was provided by HORIZON IMI2 Project AIMS2-Trials No 777394; and the HORIZON-RIA funded R2D2-MH no. 101057385.

We thank the Epi25 principal investigators, local staff from individual cohorts, and all of the patients with epilepsy who participated in the study for making possible this global collaboration and resource to advance epilepsy genetics research. This work is part of the Centers for Common Disease Genomics (CCDG) program, funded by the National Human Genome Research Institute (NHGRI) and the National Heart, Lung, and Blood Institute (NHLBI). CCDG-funded Epi25 research activities at the Broad Institute, including genomic data generation in the Broad Genomics Platform, are supported by NHGRI grant UM1 HG008895 (PIs: Eric Lander, Stacey Gabriel, Mark Daly, Sekar Kathiresan). The Genome Sequencing Program efforts were also supported by NHGRI grant 5U01HG009088-02. The content is solely the responsibility of the authors and does not necessarily represent the official views of the National Institutes of Health. We thank the Stanley Center for Psychiatric Research at the Broad Institute for supporting the genomic data generation efforts. Supplemental Epi25 phenotyping was supported by “Epi25 Clinical Phenotyping R03” National Institutes of Health (1R03NS108145-01) (PIs: Daniel Lowenstein, Samuel Berkovic).

## Supplementary Material

**Supplementary Methods**

**Supplementary Table S 1 DEX results**

**Supplementary Table S 2: Clinical Features Overview**

**Supplementary Table S 3: PBSNP AE Hyperparameters**

**Supplementary Table S 4: XGB Hyperparameter + Grid Search**

**Supplementary Table S 5: Cohort Descriptives**

**Supplementary Table S 6: Full comparison of all Models**

**Supplementary Table S 7: FN vs TN etc**

**Supplementary Table S 8: Avoidable delay estimation**

**Supplementary Figure S 1: Decision on VPA dosage**

**Supplementary Figure S 2: QC plots of Samples**

