## Supplementary Methods and Figures for "A translational multimodal machine-learning prototype predicting valproate response in epilepsy treatment"

#### *Cell model and culture conditions:*

Human induced pluripotent stem cells (hiPSCs) of two healthy donors were differentiated for 40 days as previously published [1]. HiPSC lines were maintained under feeder-free conditions on Matrigel-coated plates (0.5 mg/ml Matrigel solution in DMEM/F12 medium) in TeSR-E8 medium (Stem Cell Technologies Cat. N. 05990) and passaged twice a week (at 70-75% confluency) using 0.5 mM EDTA, with a split ratio of 1:5 or 1:6. Cells were differentiated into cortical progenitors and neurons *in vitro* as previously described. Briefly, cells were dissociated using EDTA (Day -2) and plated on Matrigel-coated plates/dishes at low confluency (20%~ 104 cells/ 5cm dish) in TeSR-E8 medium supplemented with 10µM ROCK inhibitor (Y-27632 dihydrochloride, Sigma Cat. N. Y0503). The day after, the spent medium was replaced with fresh TeSR-E8 medium without ROCK Inhibitor and cells were allowed to grow for one additional day. Two days after seeding (Day 0 of differentiation), the medium was changed to default differentiation medium (DDM), which is composed by DMEM-F12 (Thermo Fisher Cat. N. 10565042) supplemented with 1% B27 without Vitamin A (Thermo Fisher Cat. N. 12587010), 1% N-2 (Thermo Fisher Cat. N. A1370701), MEM non-Essential Amino Acids Solution (1:100 Thermo Fisher Cat. N. 11140050), Sodium Pyruvate (1:100 Thermo Fisher Cat. N. 11360070), 0.05% Bovine Albumin Fraction V (Thermo Fisher Cat. N. 15260037), $\beta$-ME (1:15000, Sigma Cat. N. M3148) and Noggin (100 ng/ml, StemCell Cat. N.78060). Medium was replenished every 2 days (until 8 days *in vitro*; DIV). Starting on 16 DIV, the medium was changed to DDM without Noggin and changed every day. The cultures used for the experiments were constantly monitored for normal progenitor cell proliferation and rosette radial glia formation in culture under phase-contrast microscope analysis. At 25 DIV, cultures were dissociated into single cells using Accutase (Termo Fisher SCIENTIFIC, Cat. N. A1110501) and plated onto Matrigel-coated plates (12-well) at a density of 50.000 cells/cm$^2$. Starting on that day, cells were maintained in a Neurobasal Medium consisting of DMEM-F12 and Neurobasal-A Medium (Thermo Fisher Cat. N. 21103049) in a 1:1 ratio, supplemented with the same factors previously used.

At DIV 40 RNA was extracted using the Qiagen RNeasy Kit following the manufacturer's protocol. RNA quality was measured using a UV-photometer (Nanodrop). All RNA extractions had a concentration above 500ng/µl and a 260/230 ratio above 2.0.

**Valproate exposure**: Cortical Neurons at 35 days of differentiation were exposed to 0.33mM - 1mM VPA (Stem Cell Technologies Cat. N. 72292) solved in DMSO. Drug concentrations were chosen based on therapeutic range and on previous experiments conducted on animal tissues [2], see Supplementary Figure S **1**.

**RNA sequencing analysis:** We sequenced two technical replicates of each of the RNA samples, i.e., a total of 12 samples adapting 3' Sequencing as published [3]. All samples passed RNA-analysis with an average read count per sample of 6,027,378 (SD 890,041). Raw count data was processed in R 4.3.3 using the DESeq2 package [4]. Genes with less than 5 reads in more than 50% of the samples or with no variance across samples were excluded. Overall, a total of 15,629 genes were included after filtering. Hierarchical clustering (Euclidean Distance, Ward.D2 algorithm) and PCA of the 2000 most variable genes showed no technical outlier, thus all samples and replicates were included (Supplementary Figure S **2**).

**Differential gene expression analysis**: Differentially expressed genes (DEX) were identified applying a linear mixed model predicting gene expression by valproate dosage (fixed effect) and Donor (random effect) adapting standard parameters of the DESeq 2 pipeline. For each donor, the log2FC and fdr-corrected p-values (Wald-test) were calculated separately based on the bayesfit function in limma [5]. A gene was considered to be DEX upon VPA exposure if the fdr was below 0.05 and the |log2FC|>1 (Supplementary Table S **1**). The resulting gene list was Gene-Set 1.

**Gene network identification:** To identify co-regulated gene-networks (modules) affected by VPA dosage we performed a weighted gene co-expression network analysis (WGCNA) as recommended [6]. In short, we quantile-normalized and log2-transformed counts and calculated the adjacency matrix (ADJ). Soft-thresholding power was set to *beta=6* (ADJ = $|cor|^{beta}$) based on the fit of the resulting network to a scale free topology ($R^2$>80%) and the visual inspection of the scree-plot of beta vs. degree. For each module Eigengene values (ME, first Principal component) were calculated. Modules with a Euclidean distance below 0.1 were merged and MEs were recalculated. To identify modules differentially regulated upon VPA treatment the same linear model approach as described above was applied, testing VPA dosage as predictor of the ME value. A Module was defined to be significantly associated with VPA if replicated in both biological replicates (i.e., fdr <0.05 and sign of log2FC identical).

#### *Data description, preprocessing and quality control steps*

##### *Reference 1000 Genome cohort (Feature embedding cohort)*

For training the SNP-based autoencoders, the 1000 Genome Phase 3 30x study (1000G) cohort [7] of 3157 samples was used. QC and filtering to SNPs present in the Discovery cohort resulted in a total number of 4.1E6 SNPs.

##### *Discovery cohort*

The Discovery cohort included data from three collection sites of the Epi25 collaborative encompassing 1334 epilepsy patients across Brussels (N= 387), Frankfurt (N= 259) and Kuopio (N= 688). Raw SNP data comprised 688,032 variants, genotyped using the GSAMD-

24v1 Illumina array across all samples. Whole exome sequencing (WES) was available for 1319 individuals. Clinical data, covering variables such as seizure type, comorbidities, gender, age, age of onset, EEG, and MRI features, was available for 1244 patients. Preprocessing and QC of data is detailed below.

**SNP data preprocessing**. SNP data was quality controlled as previously published [8]. In short, SNPs with a genotyping rate <0.95 or a HWE p-value <10E-6 were excluded as well as samples with a genotyping rate <0.95, a high rate of homozygosity (PIHAT>0.2) or with a mismatch between genetic and annotated sex. Of two or more individuals with a relatedness score about 0.2, one was included at random, and the others were excluded. After QC, SNP data was phased and subjected to imputation on the Michigan Imputation Server using the Haplotype Reference Consortium cohort (HRC r1.1 2016) as reference data. Only variants with an imputation score $R^2$>0.8 were kept. QC and imputation were done independently for each of the 3 data collection sites. After imputation the quality control pipeline was re-run. Data was pooled across the collection sites and lifted to genome build GRCh38 to match the 1000G reference dataset, resulting in a total of 1327 patients with 6.9E6 SNPs. Only SNPs also present in the reference cohort were kept, resulting in a final number of 4.1E6 SNPs.

**Whole exome sequencing preprocessing.** WES data was available for 1319 patients. VCF (Variant Call Format) files were merged using bcftools [9], followed by indexing and file normalization.

Variants were annotated using the Annovar pipeline [10] with filter for SNPs and variants with functional impact on annotated genes (option -protocol refGene, avsnp147, dbnsfp30a; and -operation g,f,f). Any variants annotated as disrupting transcriptional stop, start or splicing sites or introducing a frameshift were considered as likely gene disrupting (LGD) and processed further to calculate the PBSEQ features (see below).

**Clinical data** was available for 1244 patients including seizure/epilepsy type (for a full list of features, see Supplementary Table S 2), comorbidities (autism spectrum disorder, intellectual disabilities, psychosis), sex, age at study inclusion, age of onset, EEG components, and MRI components. Four participants were excluded due to >10% missing values. Missing data in eight sparsely populated binary variables were filled with zeros. Remaining missing values were imputed using multiple imputation by chained equations (MICE) algorithm [11] in the training and test Discovery dataset separately.

**Treatment outcome** was categorized based on VPA response, with 417 patients having undergone at least one VPA treatment regimen. For each site VPA treatment response was manually reviewed and classified as 1 = response, 2 = failure, 3 = unclassified, or 4 = unknown. Classification was done in accordance with the following criteria:

**Response:** Freedom from seizures lasting for ≥12 months which according to the treating clinician and/or the person assessing the clinical phenotype can be attributed to the ASM, e.g., after initiation or an increase of dose (and prior to initiation of another treatment for epilepsy).
**Failure:** i) seizures recurring at >50% of the pretreatment seizure frequency after the appropriate ASM has been adequately applied. Patients known to be systematically non-adherent were excluded (one seizure a year due to non-adherence has been disregarded); ii) discontinuation of VPA treatment due to any reason, including ISC (16.8%), UARs (16.1%) or both (5.1%). Reasons for discontinuation were mostly unknown (59.6%).
**Unclassified**: Information available but not classifiable (e.g., seizures recurring at <50% of the pretreatment seizure frequency).
**Unknown:** Insufficient information available (excluded).

***Independent Validation Cohort***

A newly recruited cohort comprising 202 individuals from Calgary, Canada was available for validation of the processing and prediction pipeline.
**SNP data** was available for 168 patients. Genotyping Chip was GSAMD-24v1, 686 958 variants) for 76 of the new samples, and GSAMD-24v3-0-Psych-24v1 (745 980 variants) for 92 samples. Preprocessing steps were analogously performed to the discovery cohort and independently for the two different chip types. The samples were merged and lifted to hg38, resulting in a final number of 5.4E6 SNPs and 168 patients. For Gene Set 1 and 2, 95% of the SNPs available in the Discovery cohort were available in the Validation dataset (3866 out of 4093 or 94.5% for Gene Set 1, 26 589 out of 27 870 or 95.4% for Gene Set 2). Missing SNPs in the validation data were set to major alleles for all samples.
**WES data** was available for 167 individuals. Preprocessing, QC and annotation were performed as above, analogously to the Discovery cohort.
**Clinical information** was available for 148 patients. The same variables were selected as in the Discovery cohort. Analogously to the discovery cohort, eight sparsely populated binary variables were imputed with zeros. One dimensional variable ("age of onset") had 3.4% missing values which were MICE-imputed via the learned associations from the train-Discovery cohort. No patients were removed due to >10% missing values.
**Response to VPA** was labelled as described above. A total of 19 patients were excluded due to insufficient information for classifying treatment response to VPA (class 4), resulting in 183 patients (49 responders, 134 non-responders).

#### *Autoencoder Training and PBSNP Feature Extraction*

***Generating a label agnostic genetic signature***

Selected SNPs from the 1000G cohort were used to train two separate undercomplete autoencoders (AEs), one for each Gene Set. The 1000G SNP data was split into an 80% training set and a 20% hold-out testing set. Minor allele dosage was scaled to 0, 0.5, and 1.

The starting AE Model architecture was defined as follows: The Adam optimizer was used for training, along with a LeakyReLU activation function and a mean squared error (MSE) loss function. The size of the bottleneck layer was set to 10% of the input layer size. A batch size of 32 was used during training.

A grid search approach was employed to optimize hyperparameters (Supplementary Table S 3). Three input layer dropout rates (0, 0.2, and 0.5) were evaluated, along with two activation functions for the bottleneck layer (LeakyReLU and sigmoid), and two activation functions for the output layer (linear and sigmoid). Additionally, three different symmetrical neuronal network architectures with one, two, or three layers surrounding the bottleneck layer were tested. Hidden layers of the encoder and decoder had 50%, 25%, and 12.5% of the units of the input layer respectively, if present. Overall grid search tested 36 models. Each AE model was initially trained for 50 epochs, utilizing early stopping to prevent overfitting. We employed a 3-fold cross-validation strategy to assess the model's generalizability on the training set. The best model was selected based on the MSE of decoding reconstruction.

For each Gene Set, the best model was subsequently trained for a maximum of 500 epochs with early stopping, respectively. The final model was validated on the hold out 20% 1000G test set.

***PBSNP feature extraction***

For each patient, the trained encoder models were applied to extract their compressed representation of the VPA-pathway (Gene Set 1) and metabolism pathway (Gene Set 2), i.e., the weights of the respective bottleneck nodes.

#### *Exome Sequencing based network parameters engineering*

***Graph-based feature extraction:***

This approach aims to capture topological properties that may reflect underlying genetic effects at network information level.

We used the three co-regulated gene networks as identified in the iPSC model to calculate patients' specific network parameters as follows: Based on the gene-gene correlation matrix for each network of interest we calculated graph-specific weighted parameters. This included the following: sum of connectivity (sum of lower triangle of correlation matrix), mean connectivity (mean of lower triangle of correlation matrix), average path (average of weights of all shortest paths defined by the minimum sum of weights (1-|cor|) between two genes implementing Dijkstra's Algorithm), kurtosis and skewness of the distribution of the shortest paths and max distance of the network (i.e. sum of weights of the longest of the shortest paths). To calculate unweighted network parameters, correlations (i.e., edges) were binarized. The respective threshold value was set such as 5% of the nodes remained unconnected. Similarly, the maximum number of connections, mean ego size of the nodes in the network (mean connectivity), the sum of connections, the average density and distance of the graph as well as the mean betweenness (number of shortest paths crossing through a given node) and transitivity (probability that adjacent nodes are connected) were calculated. In total 12 graph features per network were calculated (PBSEQ features). All calculations were done in the igraph package in R.

##### *Calculate patient specific PBSEQ Features*

For the patients' specific network parameters any gene (i.e., node) affected by a LGD variant was excluded from the graph by setting each correlation value to 0. Network parameters were recalculated as described above for each patient-specific correlation matrix, keeping the threshold value for binarization stable. This resulted in 12 network parameters for each module.

#### *Classifier / Recommender model*

##### *Data preparation*

**Train-test split:** The multimodal Discovery cohort dataset was randomly split into a training set for feature selection and classifier training (train-Discovery: 60%) and a hold-out test set (test-Discovery: 40%). The split was stratified by ASM response status, resulting in the presence of 47% VPA-responders in both sets. All subsequent model development and parameter tuning was done in train-Discovery only.

**Upsampling:** The mismatch between responder and non-responder classes was addressed by synthetic minority oversampling (SMOTE) as a sample balancing step. SMOTE generates synthetic samples by selecting random pairs of nearest neighbours within the minority class and creating new samples along the line segments connecting them in the feature space. A total of 17 synthetic responders were created to match the number of non-responders independently for train-Discovery and test-Discovery, corresponding to 5% of the overall sample size.

**Scaling:** All features were minmax-scaled to (0, 1) range in the train-Discovery. The scaling parameters were subsequently applied on the test-Discovery and the Validation cohort.

##### *Feature selection*

We employed all-relevant feature selection (Boruta algorithm [12]) on the training set to select promising predictors from the feature dataset. Boruta algorithm was chosen due to its ability to determine relevant features in the presence of complex relationships and interactions within the data. The threshold for feature inclusion was set at $p>0.01$. The algorithm was iteratively executed for a maximum of 200 runs to ensure comprehensive evaluation. Both confirmed and tentative features were retained during the selection process, acknowledging the potential of tentative features to contribute meaningful insights upon further investigation. The union of features obtained from 10 independent runs of the Boruta algorithm were selected to create a comprehensive feature subset. This liberal approach ensured maximum inclusion of tentative features as the subsequent gradient-boosted decision trees classifier would be able to sort out features not contributing to prediction.

##### *Classifier*

**Training:** To predict VPA response from the generated and selected features, gradient-boosted decision trees implemented by the XGBoost library [13] were employed. For hyperparameter tuning, we employed a cross-validated grid search approach with early stopping rounds set to 20 and a maximum of 1000 training iterations on train-Discovery. The model was trained to minimizing the Root Mean Squared Log Error (RMSLE) on a binary logistic objective. The grid search explored the following hyperparameter values: Subsample, N features per tree, Max depth per tree, Minimum sum of instance weight and Eta (learning rate). The full list of hyperparameters tested is available as supplement (Supplementary Table S 4).

For final training the best model was re-trained with the best hyperparameters, an increased early stopping rounds (30) and a higher maximum number of iterations (5000) for optimal performance.

**Model test and validation:** To ensure its generalizability to new data, the validity of the final predictor, including the full feature generation pipeline, was assessed in the unseen test-Discovery dataset. Subsequently, the whole pipeline including the trained prediction model was adopted for the newly recruited validation cohort.

**Model comparison:** In addition to the full model containing features from all domains, six additional models containing only a single or a combination of two feature sources (i.e., PBSEQ feat., PBSNP feat., CLIN feat.) were trained and validated to compare performances against each other. For each of these models, the procedure was analogous to the balancing, feature selection, and classifier training steps described above.

#### *Supplementary Methods references*

### Supplementary Figures

#### Supplementary Figure S 1: Decision on VPA dosage

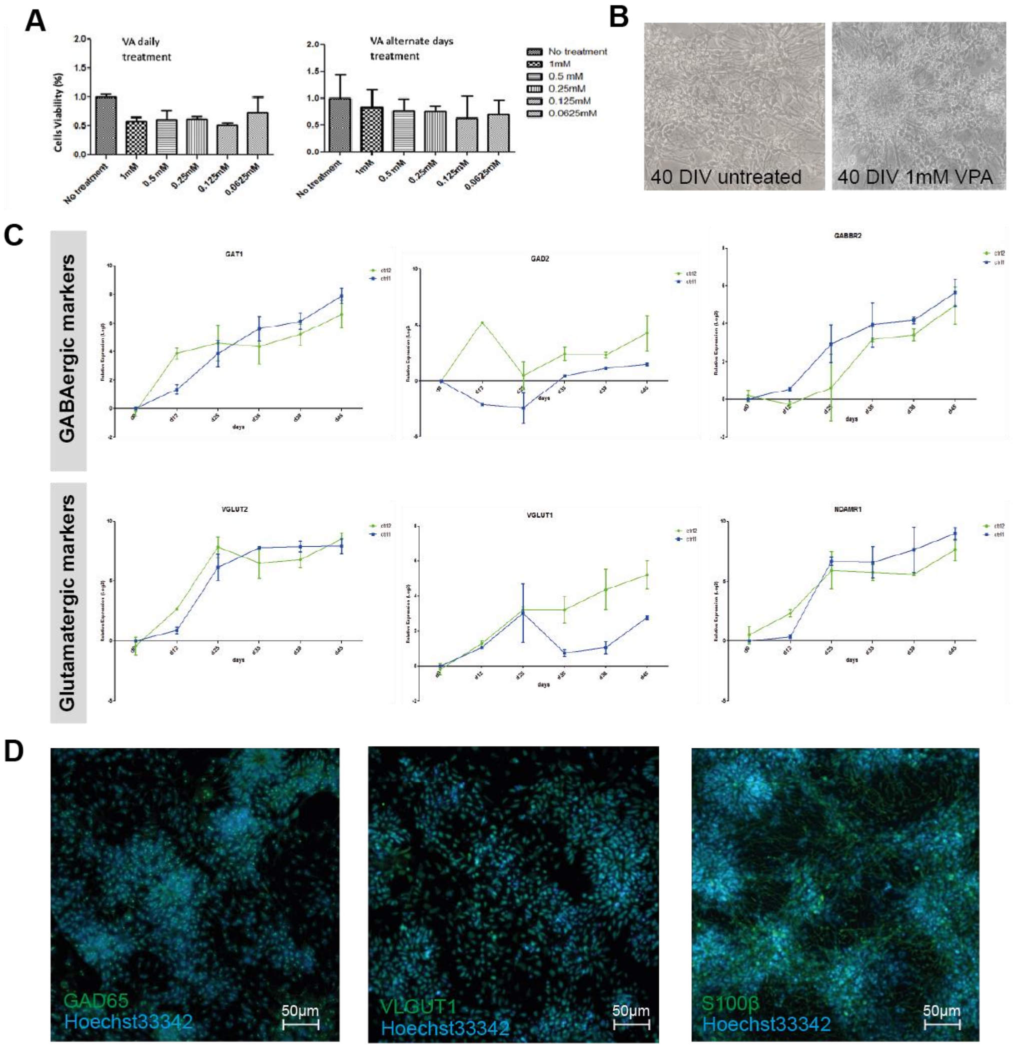

**Figure S1. A:** Cell viability assay after 40 days of differentiation shows no significant cytotoxicity across a range of VPA concentrations with daily or alternate-day treatments. **B**: Phase-contrast microscopy images show no morphological differences between untreated neurons and those treated with 1 mM VPA at 40 days *in vitro* (DIV).**C:** Longitudinal expression of GABAergic (GAT1, GAD2, GABRB2) and glutamatergic (VGLUT2, VGLUT1, NMDAR1) markers across differentiation. 2^ddCT values (GAPDH reference) are shown as mean ± SEM across thee technical replicates for the two donors (blue; green). **D:** Immunofluorescence images at 40 DIV show positive staining for GAD65 (GABAergic neurons), VGLUT1 (glutamatergic neurons), and S100β (glial cells), with nuclear staining (Hoechst 33342) confirming overall cell distribution and viability. Scale bars = 50 µm.

#### *Supplementary Figure S 2: QC plots of Samples*

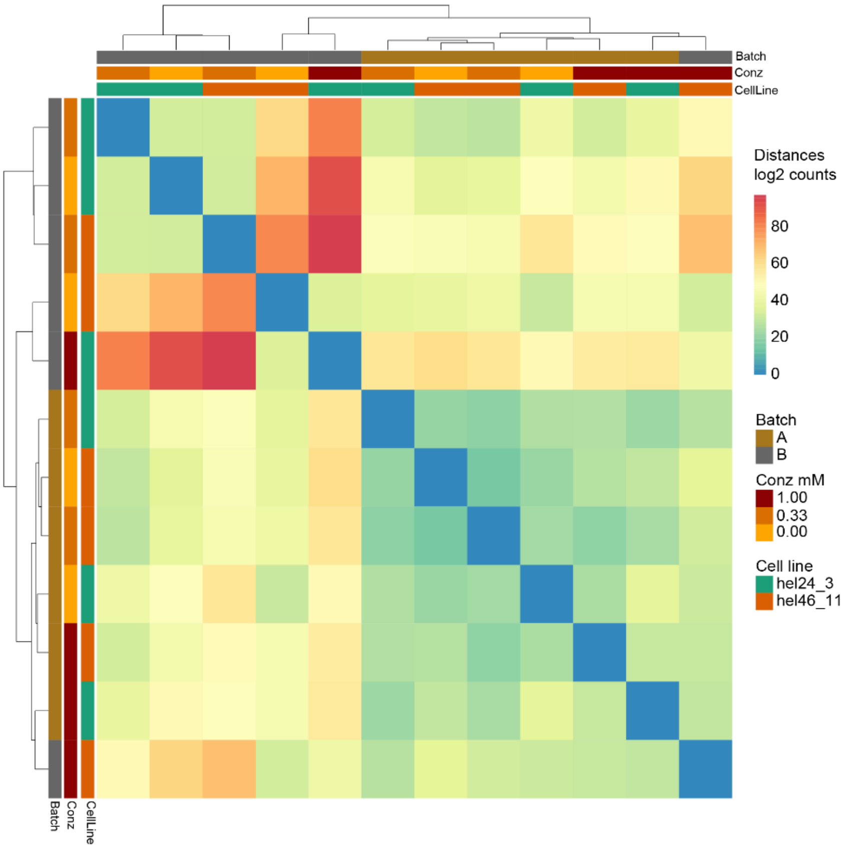

**Figure S1.** Heatmap of Euclidean distances between samples, calculated from the top 2,000 most variable genes across cell lines and technical replicates. Samples were clustered using hierarchical clustering with the Ward.D2 linkage method. The absence of strong batch-driven separation suggests good reproducibility across replicates.

### Supplementary Tables

##### Supplementary Table S 1 DEX results

Provided as Excel sheet Table_S01_VPA_DEX.xlsx

##### Supplementary Table S 2: Clinical Features Overview

Provided as Excel sheet Table_S02_CLIN_dat.xlsx

##### Supplementary Table S 3: PBSNP AE Hyperparameters

Provided as Excel sheet Table_S03_AEGridsearch.xlsx

##### Supplementary Table S 4: XGB Hyperparamter + Grid Search

Provided as Excel sheet Table_S04_XGBGridsearch.xlsx

##### Supplementary Table S 5: Cohort Descriptives

Provided as Excel sheet Table_S05_cohortDescriptives.xlsx

##### Supplementary Table S 6: Full comparison of all Models

Provided as Excel sheet Table_S06_XGBResults.xlsx

##### Supplementary Table S 7: FN vs TN etc

Provided as Excel sheet Table_S07_FNvsTP_FPvsTN.xlsx

##### Supplementary Table S 8: Avoidable delay estimation

Provided as Excel sheet Table_S08_AvoidableDelayEstimation.xlsx
